# Dynamics of Covid-19 Vaccine-Hesitancy among Primary Health Care Workers in an Urban City in India

**DOI:** 10.1101/2022.12.22.22283672

**Authors:** Syeda Sana Ali, Gowri Iyer, Hemant Mahajan, Nanda Kishore Kannuri

**Author notes:** **Corresponding author** Nanda Kishore Kannuri; Indian Institute of Public Health, Hyderabad, India. These authors have contributed equally to this work (Syeda Sana Ali, Hemant Mahajan). These authors have contributed equally to this work (Gowri Iyer, Nanda Kishore Kannuri).

## Abstract

**Background:** The health care workers (HCWs) were one of the vulnerable populations prioritized during the Covid-19 vaccination (COVISHIELD and COVAXIN) campaign. They are also the first point of contact for vaccine-related information and therefore, play a crucial role in shaping peoples’ vaccine seeking behaviour.

**Objectives:** (i) To estimate the proportion of Covid-19 vaccine hesitancy among HCWs in urban primary health care centres (UPHC) across Hyderabad; and (ii) To explore factors influencing vaccine hesitancy and vaccine acceptance in this population.

**Methods:** A cross-sectional study was conducted among 238 HCWs from 21 urban health centres in Hyderabad between June and July 2021. The prevalence of vaccine hesitancy was assessed using the questions adapted from ‘the UNICEF Guyana Covid-19 Vaccination Hesitancy Survey’. We used ‘the SAGE determinants of vaccine hesitancy’ to determine factors underlying vaccine hesitancy and acceptance.

**Results:** The prevalence of vaccine hesitancy among HCWs was 17% (12.3% - 22.2%) during the 6 months following emergency vaccine approval. ‘Self-protection’, ‘Vaccine-confidence’, and ‘Responsibility towards the general population’ were some of the reasons in favour of Covid-19 vaccination. Whereas ‘Vaccine-safety’ has emerged as the primary determinant of vaccine-hesitancy in this population. HCWs were susceptible to misinformation in the social media and in their communities, which might have shaped their opinion about the vaccines for the Covid-19.

**Conclusion:** Although the COVID-19 vaccines (COVISHIELD and COVAXIN) were approved for administration by the Drugs Controller General of India, one in every six HCWs working in the UPHCs in Hyderabad, India had either *refused or delayed* vaccinations mainly due to limited information on ‘vaccine-safety’. This highlights a critical need to address the vaccine-hesitancy among HCWs (especially during the initial phases of novel vaccine introduction), as similar behaviour of the HCWs towards novel vaccines could affect the uptake of these vaccines among the general population (which they serve).

## Introduction

A disease outbreak whether by a ‘known’ or a ‘novel’ pathogen raises concerns due to its potential effect on the health of the people. One of the tried and tested interventions to control the spread of many infectious disease outbreaks, including new diseases are ‘vaccines’ [1]. In India, to control the COVID-19 pandemic, the vaccination drive was launched on January 16^th^, 2021 [2] in a phased manner, prioritizing on vulnerable populations. On account of high risk of occupational exposure to Covid-19 [3], healthcare workers (HCWs: doctors, nurses, Accredited Social Health Activist (ASHA), etc.) were prioritized in the first-phase of the vaccination drive. The ‘COVISHIELD’ vaccine developed by the Oxford-AstraZeneca and manufactured by the Serum Institute of India, and the ‘COVAXIN’ by the Bharat Biotech were authorized for use in initial phases. Although both the vaccines were approved for administration by the Drugs Controller General of India (DCGI, the highest vaccine authority in India) and available free of cost at the government facilities and at affordable prices at the private healthcare facilities, the initial vaccine-uptake was low. By February 6^th^, 2021, only 54.7% of HCWs registered in the CoWin app were vaccinated across India [4].

The most anticipated reason for the low-vaccine coverage in India was ‘vaccine-hesitancy’. The vaccine-hesitancy is defined as ‘delay in acceptance or refusal of vaccines despite availability of vaccine services’ [5] and has been listed as the top ten global threats of 2019 [6]. The anecdotal evidence suggest the role of several contextual and individual/group influences and vaccine-specific issues underlying the vaccine-hesitancy among HCWs [7–10] such as: (i) Adverse events following vaccination/immunization (AEFIs); (ii) The spread of misinformation about the Covid-19 pandemic as well as the vaccines on social media; (iii) Fast tracking of the vaccine approval process; and (iv) The absence of quality data on vaccines safety and efficacy.

The low vaccine coverage among HCWs (especially during the initial phases of novel vaccine introduction) was worrisome. The HCWs (in Indian healthcare setting) are one of the primary sources of vaccine-related information (such as the need, safety, efficacy of vaccine etc.) for the general population and play an instrumental role to shape people’s healthcare seeking behaviour including vaccine-uptake. As HCWs are critical stakeholders to disseminate knowledge about the vaccine and frequently administer or recommend the vaccine to the general population, it is important to understand in-depth the factors underlying their (HCWs) vaccine-hesitancy to inform culturally-tailored contextual interventions. Therefore, we conducted a cross-sectional study among HCWs working in urban primary health care centres (UPHCs) in Hyderabad, (mainly caters the healthcare needs of the urban slum population) to explore the pattern of vaccine uptake and the underlying reasons for the vaccine-hesitancy.

## Materials and Methods

### Study population and design

This cross-sectional study was conducted among HCWs in UPHCs across Hyderabad from June-July 2021. An HCW in India is anyone who works in a healthcare setting including doctors, nurses, auxillary nurse midwives (ANMs), accredited social health activists (ASHAs), lab technicians, pharmacists etc. The study was conducted after seeking necessary ethics approval from the Indian Institute of Public Health, Hyderabad (Registration no. IIPHH/TRCIEC/264/2020; Dated: 28/06/2021).

There were total 85 UPHCs in Hyderabad at the time of the study. We conveniently selected 21 UPHCs (as the data collection was done amidst second wave of Covid-19 pandemic and the HCWs were scattered across Covid-19 vaccination centres) to reach a sample size of 256 (calculated based on a 95% confidence interval, anticipated proportion with vaccine hesitancy as 36.0% [11] with a relative precision of 20%). All HCWs in these 21 UPHCs who were available and who consented to participate were included in the study.

A semi-structured questionnaire were developed from the Ministry of Public Health and UNICEF Guyana Covid-19 Vaccination Hesitancy Survey [12]. It was adapted into English, Telugu and Urdu languages and pilot tested.

The study was initially designed as an online survey where a google form was created for online data collection and the link was sent to HCWs through WhatsApp. A similar methodology was used in a study in Zimbabwe and the response rate was acceptable. In India, this particular app is very popular and people check it very regularly [13]. However, due to lack of online responses, in-person data collection was carried out. During this time, the principal investigator observed that several HCWs did not have android phones and their digital literacy was very poor. Thus, the data was collected through both in-person and online. We mainly collected information on sociodemographic variables and Covid-19 vaccination status of HCWs. To assess vaccine hesitancy, we asked a very specific question: “Did you hesitate or refuse to get the Covid-19 vaccine?” To understand the perspectives on various domains such as contextual and individual/group, and vaccine-specific issues, we conducted unstructured interviews focusing on certain aspects based on ‘the SAGE determinants of vaccine hesitancy’ [5].

### Statistical Analysis

Before importing data into the statistical software, all the data was deidentified and participants were given unique IDs, which was only known to Principal investigator and co-investigators. The participants or other hospital staff outside the research group were not aware about these IDs.

The burden of vaccine-hesitancy was expressed as percentage with 95% confidence interval (calculated using binomial exact test). The qualitative data from the open-ended responses (from unstructured interviews) were arranged into themes according to ‘the SAGE determinants of vaccine-hesitancy’ to determine the factors associated with Covid-19 vaccine-hesitancy. A thematic framework was developed to unpack the factors that shaped the perceptions about the Covid-19 vaccine. The coding framework (S2 table, S3 table and S3 Fig) focused on contextual influences, individual and group influences, and vaccine specific issues.

## Results

Total 238 HCWs from 21 UPHCs participated in this study. The median (interquartile range) age of the participants was 35.0 (30.0, 40.0) years and 216 (91%) of them were females. The HCWs who participated, comprised of a diverse group of health care professional: Doctors, 8%; nurses, 15%; ASHAs, 44%; ANMs, 16% (Supp. Table 1). The ‘Others’ group, 17%, included a mix of HCWs such as multipurpose health worker, lab technicians, public health nurse, pharmacists etc. 82% of the participants were completely vaccinated with the Covid-19 vaccine. The occupation disaggregated vaccination status is given in S1 Fig. It shows that 64% doctors and 72% ASHAs in the study were vaccinated with two doses of either COVISHIELD or COVAXIN. The prevalence (95% confidence interval) of Covid-19 vaccine hesitancy among the participants was 17% (12.3% -22.2%) during the 6 months following emergency vaccine approval.

### SAGE determinants of Covid-19 vaccine hesitancy

The following factors influenced the uptake of Covid-19 vaccines and demonstrate the HCWs’ attitude towards Covid-19 vaccines and prevention (Fig 1). The vaccination status has been mentioned for those who are vaccinated with at least one dose of Covishield or Covaxin.

**Fig 1:**
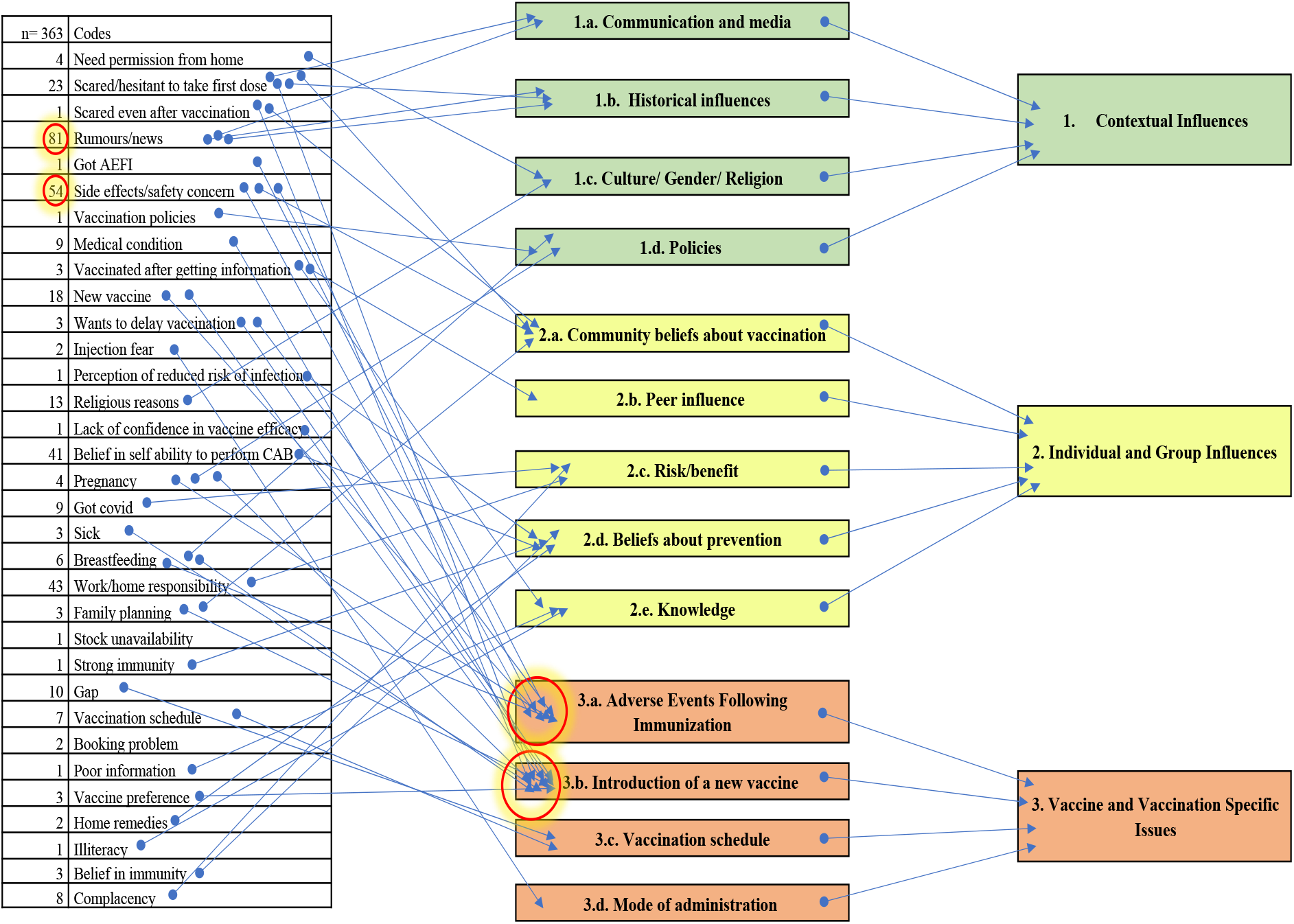
Coding framework to demonstrate the SAGE determinants of Covid-19 vaccine hesitancy among the HCWs.

#### 1. Contextual Influences

##### 1.a. Communication and media

The HCWs had delayed or refused the Covid-19 vaccines due to the spread of misinformation in the social media. Common sources of information such as news might have contributed to hesitancy.

> UID 205: *“Since the vaccine is new and there are some negative rumours spreading about the vaccine in the social media*.*”* Others, 31-35 years, male, Accountant cum clerk, vaccinated;
>
> UID 78:*”I was scared to take the 1st dose after hearing about side effects in the news”* ASHA, 26-30 years, vaccinated;

##### 1.b. Historical influences

Past attitudes towards vaccines, such as polio vaccine, seemed to have extended to the Covid-19 vaccines. There appeared to be a historical distrust in the overall health system which contributed to the suspicion that health interventions were being utilized for harmful purposes.

> UID 10: *“They are giving some other vaccines. It is birth control for Muslims”* (as narrated to them by the public) ANM, 36-40 years, vaccinated;

> UID 196: *“People spread rumours that after vaccination we will die after 2 years”* ASHA, 36-40 years, vaccinated

> UID 207: *“Vaccine will cause infertility. It will affect the reproductive system”* ASHA 31-35 years, unvaccinated

##### 1.c. Culture/ Gender/ Religion

Majority of the HCWs were of the view that men need to be prioritized for vaccination since they had an increased risk of exposure, were less likely to follow Covid-appropriate behaviour, could serve as carriers of infection and had a higher likelihood of experiencing severe disease

> UID 220: *“Men go out for work. They meet their friends without masks. Women are mostly at home. Men mingle more with friends*.*”* Nurse, male, 36-40 years, unvaccinated

Most of the female HCWs did not have the autonomy to make their own health care decisions. There was a reliance on an authoritative figure, apart from the health department and medical officer, for assurance about safety of the vaccines.

> UID 109: *“I was scared of side effects. Did not get permission from home”* ASHA, 31-35 years, unvaccinated

> UID 191: *“I got permission from home to take vaccine and that they are safe*.*”* ASHA, 26-30 years, vaccinated

The HCWs believed that vaccinations and covid appropriate behaviour along with religious beliefs provided appropriate protection.

> UID 193: *“I have taken the vaccine and I am taking precautions. I trust in God that he will protect me and my family”* ANM, 26-30 years, vaccinated

##### 1.d. Policies

By the time the data collection for this study was carried out, Covid-19 vaccines were declared safe for use in pregnancy and lactation. Despite this approval, some female HCWs who were pregnant did not take the vaccine. There was a lack of trust in the policies and also research in these vaccines. This distrust could also be because of lack of effective communication of scientific information.

#### 2. Individual and Group Influences

##### 2.a. Community beliefs about vaccination

The HCWs encountered several concerns from the public regarding vaccine safety owing to reports in the news and social media. Such misinformation may have fostered public distrust in vaccines and decreased or delayed vaccine uptake. The public was unclear about the efficacy of Covid vaccines and expected sterilizing immunity. Furthermore, there was reliance on family members for consent to vaccinate. The HCWs were able to overcome these challenges, to some extent, and were able to successfully recommend the vaccine by informing about self-vaccination. The vaccine concerns of the public/community, as narrated to the HCWs, are presented here:

> UID 171: *“I heard that infertility will occur”* Nurse, female, 26-30 years, unvaccinated

> UID 109: *“We don’t have permission from home. We are scared of side effects”* ASHA, 31-35 years, unvaccinated

> UID 68: *“Getting diseases after vaccine. People are dying. One ASHA and Anganwadi worker died. So, we explained we have taken. We are not dead. We explain then they agree to vaccinate”* 36-40 years, vaccinated

The vaccine attitude of the public was similar to that of the HCWs. Vaccine acceptance among the public was appeared to be guided by religious beliefs. There was a historical context that fostered trust in the system which develops, manufactures and delivers these vaccines. There were concerns of future unknown effects of vaccines. These factors of vaccine hesitancy among the public were the challenges faced by the HCWs during immunization campaigns.

> UID 110: *“God is there to save us. There is no need for vaccine”* Doctor, female, 26-30 years, vaccinated

> UID 189: *“The next generation will not be able to bear children. Vaccines are given to reduce population. What is the guarantee that it will protect us?”* ASHA, 36-40 years, vaccinated

> UID 102: *“There are side effects. People are getting clots. They want single dose vaccine. They want Covishield for international travel because Covaxin is not yet approved”* ASHA, 36-40 years, vaccinated

##### 2.b. Peer influence

The HCWs’ decision to vaccinate was guided by their peers. The Medical Officers were influential in generating vaccine confidence. The large-scale vaccination campaign also seemed to have motivated a HCW to vaccinate. Vaccinated HCWs demonstrated confidence in Covid-19 vaccines, as well a responsibility to raise awareness and recommend the vaccine.

> UID 160: *“To protect myself from Covid and at the same time give awareness regarding safety of vaccine and encourage my staff and people around me*.*”* Doctor, female, 36-40 years, vaccinated

> UID 69: *“We will have immunity power and then we will not get disease. Madam (MO) also explained to me”* ASHA, 46-50 years, vaccinated, vaccinated

> UID 138: *“Everyone was taking so I also took”* Attender, 56-60 years, male, vaccinated

##### 2.c. Risk/Benefit

There were variations in perceptions of risk. A nurse viewed her susceptibility to SARS-CoV-2 infection as low due to the practice of covid appropriate behaviour, which was considered superior to Covid vaccines as a method of prevention. Religious beliefs, distrust in the health system and concerns about vaccine efficacy contributed to hesitancy.

> UID 171: *“I don’t think the corona virus is dangerous to my health. For religious reasons. I don’t believe that the vaccine will stop the infection. I don’t need the vaccine because I do all the right things. I wash my hands and wear a mask and gloves. The COVID-19 vaccine is a conspiracy*.*”* Nurse, 26-30 years, female, unvaccinated

##### 2.d. Beliefs about prevention

HCWs believed that in addition to vaccines, alternative medicine and home remedies also play an important role in providing immunity from Covid-19. Belief in innate immunity were also some of the reasons for refusal to vaccinate.

> UID 143: *“Home remedies also give some immunity”* Lab technician, 36-40 years, vaccinated UID 167: *“Because I have innate immunity”* Other, 46-50 years, male, unvaccinated

##### 2.e. Knowledge

In the quote mentioned below, a nurse had confidence in the vaccine as well as knowledge that Covid-19 vaccines do not prevent infection, but are effective against reducing severe disease. This is essential, as one of the primary arguments against vaccination is the occurrence of reinfection or breakthrough infection.

> UID 124: *“Protection. Even if I get Covid after vaccination, I will get better at home only. No need for hospitalization”* Nurse, female, 36-40 years, vaccinated

#### 3. Vaccine and Vaccination Specific Issues

##### 3.a. Adverse Events Following Immunization

In addition to misinformation, fear of adverse events was one of the major contributors to Covid-19 vaccine hesitancy. The response by an ASHA captures hesitancy post vaccination. This could influence their attitude towards vaccine recommendation. The response by a doctor about no longer being concerned about adverse events implies emerging confidence in the vaccines. In this study it was apparent that there was initially a lack of trust in the research and development of Covid-19 vaccines.

> UID 161: *“Initially I was hesitant because of their side effects but now I’m not*.*”*
>
> Doctor, female, 21-25 years, vaccinated

> UID 77: *“I was scared to take the first dose. I had heard about people dying. I was scared even after taking the vaccine. I was expecting something bad to happen”* ASHA, 26-30 years, vaccinated

> UID 171: *“I am concerned about the potential side effects of the vaccine. I DO NOT think the vaccine is safe*.*”* Nurse, 26-30 years, female, unvaccinated

> UID 208: *“I was pregnant”* Nurse, 26-30 years, unvaccinated

> UID 235: *“Breastfeeding mother”* Nurse, 26-30 years, unvaccinated

##### 3.b. Introduction of a new vaccine

The concerns about a novel vaccine were heightened because of the spread of misinformation. UID 205: *“Since the vaccine is new and there are some negative rumours spreading about the vaccine in the social media*.*”* Accountant cum clerk, 31-35 years, male, vaccinated

##### 3.c. Vaccination schedule

Vaccinations were also delayed due to previous Covid infection, following which they were required to vaccinate after a gap of three months. However, previous exposure followed by delay in vaccination can also be due to a belief of having acquired immunity to Covid-19 infection.

> UID 20: *“I got Covid”* ANM, 41-45 years, unvaccinated

##### 3.d. Mode of administration

The intra muscular mode of delivery of the vaccine also appeared to contribute to vaccine-hesitancy.

> UID 167: *“Because of injection fear”* Others, 46-50 years, male, unvaccinated

## Discussion

We conducted a cross-sectional study among HCWs working in UPHCs in Hyderabad to assess the burden and underlying factors for vaccine hesitancy for the Covid-19 vaccines (COVISHIELD and COVAXIN) during the 6 months following the DCGI’s emergency vaccine approval. The prevalence of Covid-19 vaccine-hesitancy among HCWs across Hyderabad was 17% (95% CI: 12.3% -22.2%). The various factors underlying vaccine-hesitancy highlighted by the HCWs were ‘vaccine safety’ and ‘misinformation’ about the new vaccines. To our knowledge, this is the first study of its kind describing the burden and determinants of Covid-19 vaccine hesitancy among the vulnerable population in Hyderabad.

The burden of vaccine-hesitancy found in this study is considerably lower than a study conducted among HCWs in Uttar Pradesh India, which reported the prevalence of vaccine-hesitancy of 35.8% [14]. This difference could probably be due to the difference in gender distribution: 72 (28.3%) vs. 216 (91%) female participants. Another reason could be that the Uttar Pradesh study was conducted immediately after vaccine approval, whereas this study was conducted almost six months after emergency approval. The burden of hesitancy among HCWs in our study is higher when compared to the Guyana vaccine hesitancy survey, which reported a prevalence of 6% among HCWs. The reported low prevalence in that study could be due to inclusion of only 55 HCWs in the study [15]. The relatively low burden of vaccine-hesitancy among HCWs in our study compared to the Uttar Pradesh study could mean that most HCWs in UPHCs across Hyderabad had confidence in the Covid-19 vaccines as well as the government’s vaccine policy; and might have recommended the Covid-19 vaccines to the public.

We found that when the Covid-19 vaccines were given emergency approval in January 2021, some HCWs were hesitant and delayed the vaccination, while some had absolutely refused to vaccinate. Similar to previous studies [16], we observed that the ‘vaccine safety’ was one of the primary determinants of vaccine-hesitancy in this particular population. The safety issue of vaccine along with historical perceptions of childhood vaccinations such as polio, propagated by misinformation through the social media may have had an impact on early acceptance of vaccines in our study participants. Similar findings were also reported in a qualitative study in UK [17], where misinformation about the Covid-19 vaccine impacted HCWs’ attitude towards Covid-19 vaccination.

The SAGE working group have highlighted that vaccine-hesitancy can occur along a continuum from ‘complete acceptance’ to ‘complete refusal’ of vaccination [5]. We found out the similar dynamic nature of hesitancy specific to the new and rapidly developed anti-covid vaccines: COVISHIELD and COVAXIN. We captured the gradual shift in vaccine behaviour: from ‘hesitancy’ and ‘refusal’ to ‘complete acceptance’; when it was realized that the benefits of the vaccines outweigh the risks due to Covid-19. The primary reason for acceptance of vaccines was the confidence in vaccines and vaccination (to provide protection and public health responsibility of government towards community). This is similar to a study done in UK [18], where vaccinated nurses considered influenza vaccination a public health responsibility and believed in recommending the vaccine to the general population. A study to determine factors associated with vaccination behaviour among nurses towards pandemic HINI vaccine in London [19], showed that those (nurses) with a good knowledge of the disease and vaccine were more likely to recommend the vaccine to their patients. Moreover, the likelihood of recommendation was increased if the HCWs themselves had been vaccinated [18][19]. This is apparent in this study too, with 93% vaccinated HCWs were strongly agreed to recommend the Covid-19 vaccines to other people (S2 Fig).

Studies have showed that poor quality of communication can contribute to vaccine-hesitancy. The success of recommendation depends on the ability of the HCWs to recognize and understand misinformation, and effectively communicate safety and efficacy messages of the vaccines. In our study, we observed that communications about the benefits of the vaccines to the hesitant public were successful to a certain degree, with the public agreeing to vaccinate following the HCWs’ declaration of self-vaccination.

In India’s healthcare system, ASHAs and ANMs play an important role on several fronts including health promotion, vaccination, and community mobilization. The ASHAs played an important role during the Covid-19 pandemic to motivate and mobilize people to seek Covid-19 vaccination. Moreover, ASHAs have been at the forefront for delivering vaccine safety messages to the community [20]. However, sometimes even as frontline workers they did not have the autonomy or access to make the appropriate health-care decision for themselves. The Covid-19 vaccination was delayed (among ASHAs and ANMs) due to the absence of parental or spousal consent and some expectant mothers were still (at the time of interview) unvaccinated, despite evidence of vaccine safety [21] and effectiveness [22]. In our study it was apparent that there was initially a lack of trust in the research and development of Covid-19 vaccines. It was also apparent that the HCWs were concerned about the use of these vaccines during pregnancy and lactation. These findings are similar to a study done in Germany which studied the acceptance of Covid-19 vaccines among pregnant and breastfeeding women [23]. The gender related barriers faced by female HCWs can be overcome by community engagement [24] and sharing scientific data about the benefits of vaccination for pregnant women and neonates [22] to their families to achieve equitable vaccine coverage.

The confidence of HCWs in vaccination could be improved by providing them with correct and timely information. In this study, the HCWs reported that they periodically received vaccine-related information from the routine office meetings. The information provided in such meetings should be verified for the quality and content to ensure that it correctly addresses vaccine-related issues and other specific vaccine related concerns (as discussed earlier). Further studies are required to understand the impact of periodic training and sharing of information to HCWs.

We also observed that the concerns of the HCWs in this study regarding Covid-19 vaccines were similar to those of the public. Since many HCWs belongs to the community which they serve, social influence cannot be ruled out. Evidence [25], shows that vaccine hesitancy among public could contribute to hesitancy among HCWs. In a qualitative study to determine vaccine hesitancy among HCWs across Europe [26], HCWs were influenced by their patients and social media. Since vaccine-hesitancy is an attitude, mere improvement in knowledge will not translate to positive attitude, improved recommendations and increased vaccine coverage.

The frequency and intensity of outbreaks by novel pathogens is predicted to increase in the future [27]. In such a situation, hesitancy to rapidly developed new vaccines is to be anticipated. Furthermore, in such events it is expected that HCWs will be one of the most vulnerable yet valuable population, and will therefore be the first to be protected using newly developed vaccines. Therefore, developing trust and transparency in vaccine research and vaccine policies among HCWs as well as the general population is essential to promote vaccine confidence. Anecdotal evidence suggests that historical concerns about the adverse effects of vaccines have extended to and affected the acceptance of Covid-19 vaccines [28]. The concerns, which arose during previous vaccination campaigns, such as polio immunization for children, might have extended to Covid-19 vaccination campaigns for adults [29]. It is possible that such lack of vaccine-confidence may extend to future pandemic vaccines also.

As part of pandemic preparedness, global partnerships like CEPI [30] aim to accelerate the development of pandemic vaccines. Such an initiative requires the support of HCWs to ensure vaccine confidence among the public towards these rapidly developed new vaccines. During a pandemic it is crucial to not just increase vaccination-coverage (the proportion of population who receive the lifesaving vaccine) but also the rate of vaccine-coverage (the speed at which they receive the vaccine). There is an urgent need to address the reasons for hesitancy towards pandemic vaccines among HCWs, as this delay and refusal could extend to the public. Lessons can be learnt from the vaccination programs for childhood vaccination, which is ongoing in India since last 70 years. On very similar line, the overall health infrastructure and health system can be developed to enable an environment for adult vaccination in this and upcoming pandemics. With new vaccines emerging for Covid-19 and prospective pandemic vaccines, HCWs will find it challenging to adapt to the changing vaccine environment and therefore, the need for vaccination efforts needs to be emphasized. Behaviour models such as the ‘health-belief model’ can be used to predict vaccine-hesitancy and develop tailored strategies [31].

The strengths of the study include: In-person interviews were conducted for most of the participants by a trained interviewer using a validated questionnaire and vaccine-hesitancy framework. Limitations of the study are: (i) Vaccination status was self-reported and we could not confirm the actual vaccine administration of participants. Therefore, the possibility of social desirability bias cannot be completely excluded. Nevertheless, it is very unlikely that participants might have forgotten or hidden their vaccination status considering the situation of the pandemic. (ii) Since we used convenience sampling to recruit participants amidst the second wave of COVID-19 in India, caution should be exercised while generalizing the findings to the general population or the rest of the country. (iii) There were some concerns about Covid-19 vaccines even among the vaccinated HCWs. This aspect of vaccine hesitancy could not be captured. Despite these limitations, this study provides us with a window into some of the concerns regarding the Covid-19 vaccine-hesitancy among HCWs serving the urban marginalized communities in Hyderabad and the challenges faced by them.

## Conclusion

In this study, the prevalence of Covid-19 vaccine hesitancy was 17%, which mean that we found that every sixth HCWs across UPHCs in Hyderabad was hesitant to receive the newly developed Covid-19 vaccines during the 6 months following administrative approval. The factors contributing to hesitancy in this particular population were: distrust in the pandemic vaccine research and development, distrust in vaccine policies, fear of AEFI and misinformation about the pandemic and the vaccines. Further research is required for in-depth exploration of these factors to inform context-specific interventions.

## Supporting information

Supplemental Table 1. Descriptive Characteristics of Study Participants

Supplemental Table 2. Coding Framework

Supplemental Table 3. Count and Percentage of each Quote

Supplemental Fig 1. Occupation of HealthCare Workers

Supplemental Figure 2. Attitude of Healthcare Workers towards Covid-19 Vaccination

## Data Availability

Data cannot be shared publicly because of Data privacy policies of Indian Institute of Public Health, Hyderabad, India. The Data are available from the Indian Institute of Public Health, Hyderabad from the Institutional Data Access / Ethics Committee; contact via email to Dr Nanda Kishore Kannuri for researchers who meet the criteria for access to confidential data.

## Authors’ Contribution

SSA and GI formulated the research question. All authors contributed to the design of the study. SSA, GI, and NKK prepared the protocol. GI and NKK supervised the study. SSA and HM curated the data, conducted the formal data analysis and wrote the first draft of the paper. All authors edited the subsequent drafts and approved the final version of the paper.

## Acknowledgment

We thank Konkati Shiva Prasad for his assistance in translation of the questionnaire into Telugu.

## References

[1] Savulescu C, Jiménez-Jorge S, De Mateo S, et al. Using surveillance data to estimate pandemic vaccine effectiveness against laboratory confirmed influenza A(H1N1)2009 infection: Two case-control studies, Spain, season 2009-2010. BMC Public Health 2011; 11: 1–9.

[2] India rolls out the world’s largest COVID-19 vaccination drive, https://www.who.int/india/news/feature-stories/detail/india-rolls-out-the-world-s-largest-covid-19-vaccination-drive (accessed 20 March 2022).

[3] Gholami M, Fawad I, Shadan S, et al. COVID-19 and healthcare workers: A systematic review and meta-analysis. Int J Infect Dis 2021; 104: 335–346.

[4] Covid-19: India vaccinates 54.7% healthcare workers registered on CoWin platform | India News - Times of India, https://timesofindia.indiatimes.com/india/covid-19-india-vaccinates-54-7-healthcare-workers-registered-on-cowin-platform/articleshow/80725869.cms (accessed 19 February 2021).

[5] WHO. Report of the Sage Working Group on. 2014; 64.

[6] Ten threats to global health in 2019, https://www.who.int/news-room/spotlight/ten-threats-to-global-health-in-2019 (accessed 26 February 2021).

[7] Ashok N, Krishnamurthy K, Singh K, et al. High COVID-19 Vaccine Hesitancy Among Healthcare Workers: Should Such a Trend Require Closer Attention by Policymakers? Cureus; 13. Epub ahead of print 15 September 2021. DOI: 10.7759/CUREUS.17990.

[8] Indians hesitant to get Covaxin, Covishield Covid-19 vaccines — Quartz India, https://qz.com/india/1959402/indians-hesitant-to-get-covaxin-covishield-covid-19-vaccines/ (accessed 21 February 2021).

[9] Hesitancy to take Covid-19 vaccine reason for low turnout in India - The Hindu BusinessLine, https://www.thehindubusinessline.com/news/covid-19-vaccine-hesitancy-responsible-for-low-turnout-in-india/article33615039.ece (accessed 18 February 2021).

[10] Coronavirus | Covaxin uptake less than 1% in 12 States, UTs - The Hindu, https://www.thehindu.com/news/national/coronavirus-covaxin-uptake-less-than-1-in-12-states-uts/article33906644.ece (accessed 11 September 2022).

[11] As PM Modi, Ministers take COVID-19 jab & vaccination begins at private hospitals, vaccine hesitancy drops to 36%, https://www.localcircles.com/a/press/page/co-win-site-issues-survey#.YxoBF3ZBy3A (2022).

[12] Survey V. Ministry of Health (Maternal and child Health Department) in Partnership with UNICEF - COVID-19 Vaccine Survey. 2021; 1–8.

[13] Mundagowa PT, Tozivepi SN, Chiyaka ET, et al. Assessment of COVID-19 vaccine hesitancy among Zimbabweans: A rapid national survey. PLoS One; 17. Epub ahead of print 1 April 2022. DOI: 10.1371/JOURNAL.PONE.0266724.

[14] Singh AK, Kumari R, Singh S, et al. The dilemma of covid-19 vaccination among health care workers (Hcws) of uttar pradesh. Indian J Community Heal 2021; 33: 319– 324.

[15] 2020 https://www.surveymonkey.com/r/KAP_GY.AJ. Ministry of Public Health, in Partnership with UNICEF - KAP Survey on COVID 19-Response.

[16] Vaccine hesitancy: A growing challenge for immunization programmes, https://www.who.int/news/item/18-08-2015-vaccine-hesitancy-a-growing-challenge-for-immunization-programmes (accessed 3 June 2022).

[17] Manby L, Dowrick A, Karia A, et al. Healthcare workers’ perceptions and attitudes towards the UK’s COVID-19 vaccination programme: a rapid qualitative appraisal. BMJ Open 2022; 12: e051775.

[18] Paterson P, Meurice F, Stanberry LR, et al. Vaccine hesitancy and healthcare providers. Vaccine 2016; 34: 6700–6706.

[19] Zhang J, While AE, Norman IJ. Nurses’ vaccination against pandemic H1N1 influenza and their knowledge and other factors. Vaccine 2012; 30: 4813–4819.

[20] The ASHA Women Delivering COVID-19 Vaccines in India – The Wire Science, https://science.thewire.in/health/the-asha-women-delivering-covid-19-vaccines-in-india/ (accessed 3 April 2022).

[21] Blakeway H, Prasad S, Kalafat E, et al. COVID-19 vaccination during pregnancy: coverage and safety. Am J Obstet Gynecol 2022; 226: 236.e1-236.e14.

[22] Dagan N, Barda N, Biron-Shental T, et al. Effectiveness of the BNT162b2 mRNA COVID-19 vaccine in pregnancy. Nat Med 2021 2710 2021; 27: 1693–1695.

[23] Schaal NK, Zöllkau J, Hepp · Philip, et al. Pregnant and breastfeeding women’s attitudes and fears regarding the COVID-19 vaccination. Arch Gynecol Obstet 2022; 306: 365–372.

[24] Building Vaccine Confidence Through Community Engagement. DOI: 10.1111/disa.12161.

[25] Nzaji MK, Ngombe LK, Mwamba GN, et al. Acceptability of Vaccination Against COVID-19 Among Healthcare Workers in the Democratic Republic of the Congo. Pragmatic Obs Res 2020; 11: 103.

[26] Karafillakis E, Dinca I, Apfel F, et al. Vaccine hesitancy among healthcare workers and their patients in Europe. 2016.

[27] Marani M, Katul GG, Pan WK, et al. Intensity and frequency of extreme novel epidemics. Proc Natl Acad Sci U S A; 118. Epub ahead of print 31 August 2021. DOI: 10.1073/PNAS.2105482118.

[28] History Does Repeat: Pandemic Vaccine Uproar Is Nothing New, https://www.webmd.com/vaccines/covid-19-vaccine/news/20211014/vaccine-opposition-not-new (accessed 25 October 2022).

[29] Yasmin F, Asghar W, Babar MS, et al. Acceptance Rates and Beliefs toward COVID-19 Vaccination among the General Population of Pakistan: A Cross-Sectional Survey. Am J Trop Med Hyg 2021; 105: 1230.

[30] CEPI | New Vaccines For A Safer World, https://cepi.net/ (accessed 4 June 2022).

[31] Limbu YB, Gautam RK, Pham L. The Health Belief Model Applied to COVID-19 Vaccine Hesitancy: A Systematic Review. Epub ahead of print 2022. DOI: 10.3390/vaccines10060973.

