## Supplemental Table 1. Descriptive Characteristics of Study Participants for "Dynamics of Covid-19 Vaccine-Hesitancy among Primary Health Care Workers in an Urban City in India"

### S1 Table: Descriptive statistics

| Variables | Categories | Descriptive Statistics (n=238) |
|---|---|---|
| Age in years, Q2 (Q2, Q3) | | 35 (30,40) |
| Gender, n (%) | Male | 22 (9) |
| | Female | 216 (91) |
| Marital status, n (%) | Married | 214 (89.9) |
| | Unmarried | 24 (10) |
| Education, n (%) | Completed High school | 61 (68) |
| | Graduate | 162 (26) |
| | Postgraduate | 15 (6) |
| Occupation, n (%) | Doctor | 19 (7.98) |
| | Nurse | 34 (14.28) |
| | Health worker | 4 (1.68) |
| | ASHA | 103 (43.27) |
| | ANM | 38 (15.96) |
| | Others | 40 (16.8) |
| Vaccination Status, n (%) | $1^{st}$ dose | 202 (85) |
| | $2^{nd}$ dose | 166 (70) |
| Vaccine hesitancy, n (%) | Yes | 40 (17) |
| | No | 198 (83) |
| What is your main source of information? (n= 809) * | Newspaper | 146 (18%) |
| | Government website | 59 (7.3%) |
| | Facebook | 38 (4.7%) |
| | WhatsApp | 63 (7.8%) |
| | Twitter | 27 (3.3%) |
| | YouTube | 58 (7.16%) |
| | Friends | 69 (8.5%) |
| | Colleagues | 183 (22.6%) |
| | Others | 166 (20.5%) |
| Do you believe that there are other (better) ways to protect yourself from covid-19 other than vaccines? (n= 411) * | Home remedies | 128 (31.4%) |
| | Alternative medicine | 81 (19.7%) |
| | Natural infection | 38 (9.24%) |
| | Others | 164 (39.9%) |

*For the main source of information and alternate methods of prevention, we received multiple response
