## Supplemental Table 2. Coding Framework for "Dynamics of Covid-19 Vaccine-Hesitancy among Primary Health Care Workers in an Urban City in India"

**S2 Table: Coding Framework**

| Respondent | Vaccination status* | Theme | Subtheme | Code | Examples |
|---|---|---|---|---|---|
| Accountant cum Clerk UID 205, 31-35 years, male | Vaccinated | 1. Contextual Influences | 1.a. Communication and media | Rumours in social media | “Since the vaccine is new and there are some negative rumours spreading about the vaccine in the social media” |
| ASHA UID 78, 26-30 years | Vaccinated | | | AEFI in news | "I was scared to take the 1st dose after hearing about side effects in the news” |
| ANM UID 10, 36-40 years | Vaccinated | | 1.b. Historical influence | Rumours | “They are giving some other vaccines. It is birth control for Muslims" (as narrated to them by the public) |
| ASHA UID 196, 36-40 years | Vaccinated | | | Rumours | “People spread rumours that after vaccination we will die after 2 years” |
| ASHA UID 109, 31-35 years | Unvaccinated | | 1.c. Culture/ Gender/ Religion | Lack of autonomy | “I was scared of side effects. Did not get permission from home” |
| ANM UID 193, 26-30 years | Vaccinated | | | Religious beliefs | “I have taken the vaccine and I am taking precautions. I trust in God that he will protect me and my family” |
| Nurse UID 171, female, 26-30 years | Unvaccinated | 2. Individual and Group Influences | 2.a. Community beliefs about vaccination (as narrated to the HCWs by the public) | Rumours | “I heard that infertility will occur” |
| ASHA UID 109, 31-35 years | Unvaccinated | | | Lack of autonomy | “We don’t have permission from home. We are scared of side effects” |
| ASHA UID 102, 36-40 years | Vaccinated | | | Vaccine preference | “There are side effects. People are getting clots. They want single dose vaccine. They want Covishield for international travel because Covaxin is not yet approved” |
| ASHA UID 69, 46-50 years | Vaccinated | | 2.b. Peer influence | Peer influence | “We will have immunity power and then we will not get disease. Madam (MO) also explained to me” |

| | | | | | |
|---|---|---|---|---|---|
| Nurse UID 171, 26-30 years, female | Unvaccinated | | 2.c. Risk/benefit | Lack of confidence in vaccine | “I don’t think the corona virus is dangerous to my health. For religious reasons. I don’t believe that the vaccine will stop the infection. I don’t need the vaccine because I do all the right things. I wash my hands and wear a mask and gloves. The COVID-19 vaccine is a conspiracy.” |
| UID 143, lab technician, 36-40 years | Vaccinated | | 2.d. Beliefs about prevention | Home remedies | “Home remedies also give some immunity” |
| Others UID 167, 46-50 years, male | Unvaccinated | | | Belief in immunity | "Because I have innate immunity" |
| Attender, UID 188, male, 26-30 years | | | 2.e. Knowledge | Source of knowledge | “Newspaper, Government website, Friends, Colleagues, Doctor, elders at home” |
| ASHA UID 77, 26-30 years | Vaccinated | 3. Vaccine and Vaccination Specific Issues | 3.a. Adverse Events Following Immunization | Rumours, Safety concern | “I was scared to take the first dose. I had heard about people dying. I was scared even after taking the vaccine. I was expecting something bad to happen” |
| Nurse UID 171, 26-30 years, female | Unvaccinated | | | Safety concern | “I am concerned about the potential side effects of the vaccine. I DO NOT think the vaccine is safe.” |
| Nurse UID 208, 26-30 years | Unvaccinated | | | Safety concern during pregnancy | "I was pregnant" |
| Nurse UID 235, 26-30 years | Unvaccinated | | | Safety concern during lactation, early vaccine policies | "Breastfeeding mother" |
| UID 205, 31-35 years, Accountant cum clerk, male. | Vaccinated | | 3.b. Introduction of a new vaccine | New vaccine | "Since the vaccine is new and there are some negative rumours spreading about the vaccine in the social media." |
| ANM UID 20, 41-45 years | Unvaccinated | | 3.c. Vaccination schedule | Vaccination schedule | “I got covid” |

| | | | | | |
|---|---|---|---|---|---|
| UID 167, 46-50 years, male | Unvaccinated | | 3.d. Mode of administration | Fear of injection | "Because of injection fear" |
| Footnote: * At least 1 dose of either Covaxin or Covishield | | | | | |
