## Supplemental Table 3. Count and Percentage of each Quote for "Dynamics of Covid-19 Vaccine-Hesitancy among Primary Health Care Workers in an Urban City in India"

**S3 Table: Count and percentage of each quote**

| Codes | n= 363 | (%) |
|---|---|---|
| Need permission from home | 4 | 1.1 |
| Scared/hesitant to take first dose | 23 | 6.34 |
| Scared even after vaccination | 1 | 0.28 |
| Rumours/news | 81 | 22.31 |
| Got AEFI | 1 | 0.28 |
| Side effects/Safety concern | 54 | 14.88 |
| Vaccination policies | 1 | 0.28 |
| Medical condition | 9 | 2.48 |
| Vaccinated after getting information | 3 | 0.83 |
| New vaccine | 18 | 4.96 |
| Wants to delay vaccination | 3 | 0.83 |
| Injection fear | 2 | 0.55 |
| Perception of reduced risk of infection | 1 | 0.28 |
| Religious reasons | 13 | 3.58 |
| Lack of confidence in vaccine efficacy | 1 | 0.28 |
| Belief in self ability to perform CAB | 41 | 11.29 |
| Pregnancy | 4 | 1.1 |
| Got covid | 9 | 2.48 |
| Sick | 3 | 0.83 |
| Breastfeeding | 6 | 1.65 |
| Work/home responsibility | 43 | 11.85 |
| Family planning | 3 | 0.83 |
| Stock unavailability | 1 | 0.28 |
| Strong immunity | 1 | 0.28 |
| Gap | 10 | 2.75 |
| Vaccination schedule | 7 | 1.93 |
| Booking problem | 2 | 0.55 |
| Poor information | 1 | 0.28 |
| Vaccine preference | 3 | 0.83 |
| Home remedies | 2 | 0.55 |
| Illiteracy | 1 | 0.28 |
| Belief in immunity | 3 | 0.83 |
| Complacency | 8 | 2.2 |
| Footnote: n is the total number of codes. Some responses have been placed under more than one code. | | |
