## Supplementary figures and images for "Dynamics of Covid-19 Vaccine-Hesitancy among Primary Health Care Workers in an Urban City in India"

### Supplemental Fig 1. Occupation of HealthCare Workers

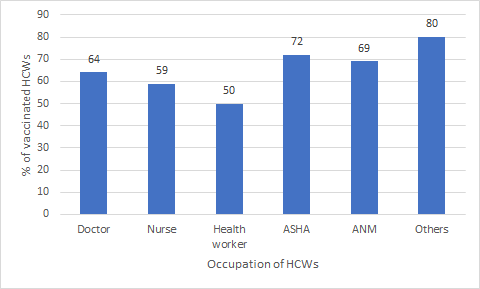

### Supplemental Figure 2. Attitude of Healthcare Workers towards Covid-19 Vaccination

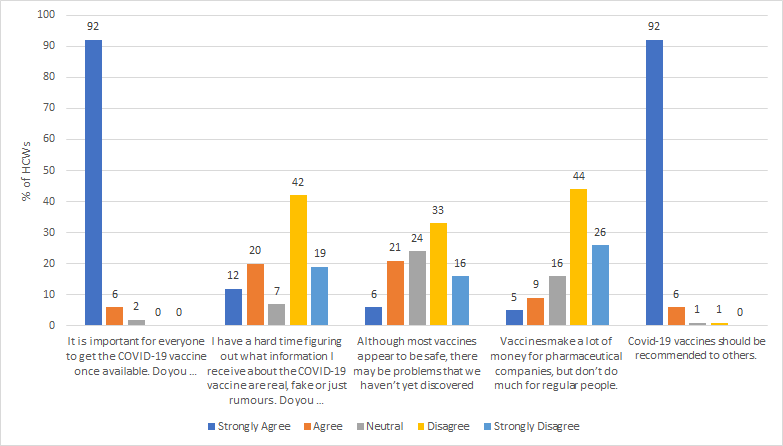
